# Segmentation and Measurements of Major Blood Vessels from CT Pulmonary Angiography Using Deep Learning

**DOI:** 10.1101/2024.11.19.24317538

**Authors:** Ali Teymur Kahraman, Tomas Fröding, Dimitris Toumpanakis, Christian Jamtheim Gustafsson, Tobias Sjöblom

## Abstract

**Background:** Measuring major blood vessels from CT pulmonary angiography examinations (CTPAs) to assess cardiovascular diseases has the potential to improve overall patient outcomes. However, this process is time-consuming and prone to errors. Deep learning (DL) approaches offer the potential to enhance accuracy, speed, and consistency.

**Objectives:** To develop and train a deep learning-based algorithm capable of automatically and accurately segmenting and measuring major blood vessels in CTPAs.

**Methods:** Seven hundred CTPAs from 652 patients were retrospectively collected at a single center. The dataset was split into two subsets, one for training and cross-validation (n = 490) and one for assessing model performance (n = 210). The segmentation masks for the descending aorta (DAo), ascending aorta (AAo), and pulmonary trunk (PT) were generated by our previously developed segmentation model and were quantitatively validated by two radiologists. These validated masks were subsequently used as ground truth for model training. An U-Net deep learning model was created using the nnU-Net framework and trained on 490 CTPAs with 5-fold cross-validation. Following the training, the model was applied to volumes of interest in the images to generate a pool of candidate regions containing potential vessels. A vessel detection algorithm was developed and used on the candidate pool to identify vessels followed by measurement. The final model was evaluated on 210 and 47 CTPAs from internal and external datasets, respectively.

**Results:** Assessing model segmentation performance on the internal evaluation set, the median Dice scores were 0.95 for the DAo, 0.96 for the AAo, and 0.95 for the PT. The model measurements showed a strong correlation with those made by the radiologist, with Pearson’s r values of 0.91 for image noise, 0.98 for intravenous contrast concentration in the PT, 0.93 for AAo diameter, and 0.55 for PT diameter (*P* < .001). Additionally, the AAo segmentation (median Dice score 0.94) and the PT diameter measurement (r = 0.77) were evaluated in two external datasets.

**Conclusions:** The fully automated, deep learning-based algorithm accurately segmented and measured major blood vessels in real-world CTPAs.

## Introduction

Cardiovascular disease (CVD) is the leading cause of morbidity and mortality worldwide [1]. Identification and accurate measurement of major blood vessels within the thoracic cavity is essential for radiologists in the diagnosis and assessment of CVDs [2–4]. Computed tomography (CT) pulmonary angiography (CTPA) is the gold standard for diagnosing pulmonary embolism (PE) and involves the use of an intravenous (IV) contrast agent to enhance the visualization of pulmonary arteries [5,6]. Additionally, CTPA enhances the visibility of other major blood vessels in the chest cavity, enabling radiologists to assess CVDs more effectively [7,8]. Several CVDs, such as chronic thromboembolic pulmonary hypertension (CTEPH), pulmonary arterial hypertension (PAH), and aortic aneurysms, can be diagnosed or suspected based on the presence of enlarged pulmonary arteries or an enlarged ascending aorta [9–11]. Therefore, a fully automated solution has the potential to improve radiologists’ workflow efficiency and accuracy and enhance overall patient outcomes.

Existing automatic methods for segmenting and measuring major blood vessels in the chest cavity often rely on traditional image processing and analysis techniques [12–14]. While deep learning-based solutions have shown promise [15–17], many existing datasets have limitations, including small sample sizes and a lack of representative artifacts and co-morbidities. To address these challenges, we propose training and testing the state-of-the-art semantic segmentation deep learning framework, the no-new-U-Net (nnU-Net) [18], on a large and diverse dataset of CTPA examinations. This dataset is designed to accurately reflect the range of imaging conditions encountered in clinical practice [19]. The nnU-Net framework represents a significant advancement in U-Net model training, automating the process of hyperparameter optimization and model selection [18]. In this study, we developed a deep learning model utilizing the nnU-Net architecture to automate the segmentation and measurement of major blood vessels within the thoracic cavity, specifically the descending aorta (DAo), ascending aorta (AAo), and pulmonary trunk (PT), in routine clinical contrast-enhanced CTPAs.

## Materials and Methods

### Internal dataset

We retrospectively collected a dataset of 700 non-ECG-gated CTPA examinations performed between 2014 and 2018 at a single institution (Nyköping Hospital, Sweden), with approval from the Swedish Ethical Review Authority (EPN Uppsala Dnr 2015/023 and 2015/023/1) for the collection and analysis of CTPA examinations [19]. The CTPAs were routine clinical examinations, exported sequentially from the history list in the institution’s Picture Archiving and Communication System (PACS) in Digital Imaging and Communications in Medicine (DICOM) format, with all personal identifiers removed from the headers using the Dicom2USB hardware solution (www.dicom2usb.com).

### CT Acquisition Protocols

Five CT scanners were used to acquire the CTPAs: Brilliance 64, Ingenuity Core, and Ingenuity CT (Philips, Netherlands); LightSpeed VCT (GE Healthcare, USA); and Somatom Definition Flash (Siemens, Germany). Scans were performed using a bolus tracking method with the region of interest in the pulmonary trunk, utilizing pixel spacing of 0.59–0.98 mm, tube voltages of 80– 120 kV, and slice thicknesses of 0.625–2.0 mm. The contrast medium (Omnipaque 350 mg I/ml, GE Healthcare) was administered in doses ranging from 20 to 114 ml (mean 62 ml) at injection rates of 2.4 to 6.1 ml/s (mean 3.6 ml/s). Secondary reconstructions with a 2.0 mm slice thickness were applied to all examinations in the internal dataset. All annotations and measurements were performed in the axial plane, providing standardized data for model development and inference.

### Data annotation

In previous work, we developed a fully automated deterministic algorithm for segmenting and measuring major vessels in CTPA examinations [19]. In this study, we utilized the previously developed deterministic solution to generate 2D segmentation masks, which serve as the ground truth for the major blood vessels in the thoracic cavity: the descending aorta (DAo), ascending aorta (AAo), and pulmonary trunk (PT). These ground truth segmentation masks were validated by two radiologists, DT and TF, with 7 and 17 years of experience in chest radiology, respectively. A single 2D segmentation mask for each vessel was obtained from each CTPA volume at a specific anatomical location, either at the carina of the trachea or the level of the pulmonary trunk. This automated process successfully generated 2D segmentation masks for major blood vessels in 596 out of 700 CTPAs, with validation by the radiologists, resulting in a total of 1,788 validated masks. However, the remaining 104 CTPAs required manual annotation, as they could not be processed automatically. This manual annotation produced an additional 312 2D segmentation masks for the three major blood vessels, using the open-source software Medical Imaging Interaction Toolkit (MITK) [20]. The manual annotations for these 104 CTPAs were also validated by DT and TF to ensure consistency and accuracy. The manual (DT and TF) ground truth measurements of major blood vessels included the diameter of the AAo and the PT, the IV contrast concentration in the PT (mean HU value in 2 cm^2^ ROI) and the image noise (standard deviation of HU in 1 cm^2^ ROI in the DAo), as reported [19].

### Examination quality of CTPA volume images

The examination quality metrics and methodologies applied were developed and validated in [19], where a comprehensive analysis was conducted on the CTPA images to establish robust quality scoring criteria. In short, for each CTPA examination the radiologist assessed five image quality parameters that affect the evaluation for PE: motion artifacts, streak artifacts, IV contrast concentration in the PT, parenchymal disease and image noise. Each parameter was scored, resulting in classification of the quality of the CTPA as good, acceptable or inferior. For detailed information on the original formulation and calculation of the quality scores, please refer to the supplementary information of [19].

### External datasets

Two publicly available datasets were used to externally evaluate the proposed algorithm: the Segmentation of Thoracic Organs at Risk in CT images (SegTHOR) [21] and the Ferdowsi University of Mashhad’s PE dataset (FUMPE) [22]. The SegTHOR training set contains 40 CT scans with and without IV contrast, all with manual segmentation of the aorta. For the external evaluation of ascending aorta segmentation, 12 CT scans with IV contrast were selected from the 40 CT scans, while the remaining 28 CTs without IV contrast were excluded. The FUMPE dataset contains 35 CTPAs with the diameter of the pulmonary trunk (PT) annotated by radiologists, which were used for the external evaluation. An overview of our internal and external datasets is shown in Figure 1.

**Figure 1.**
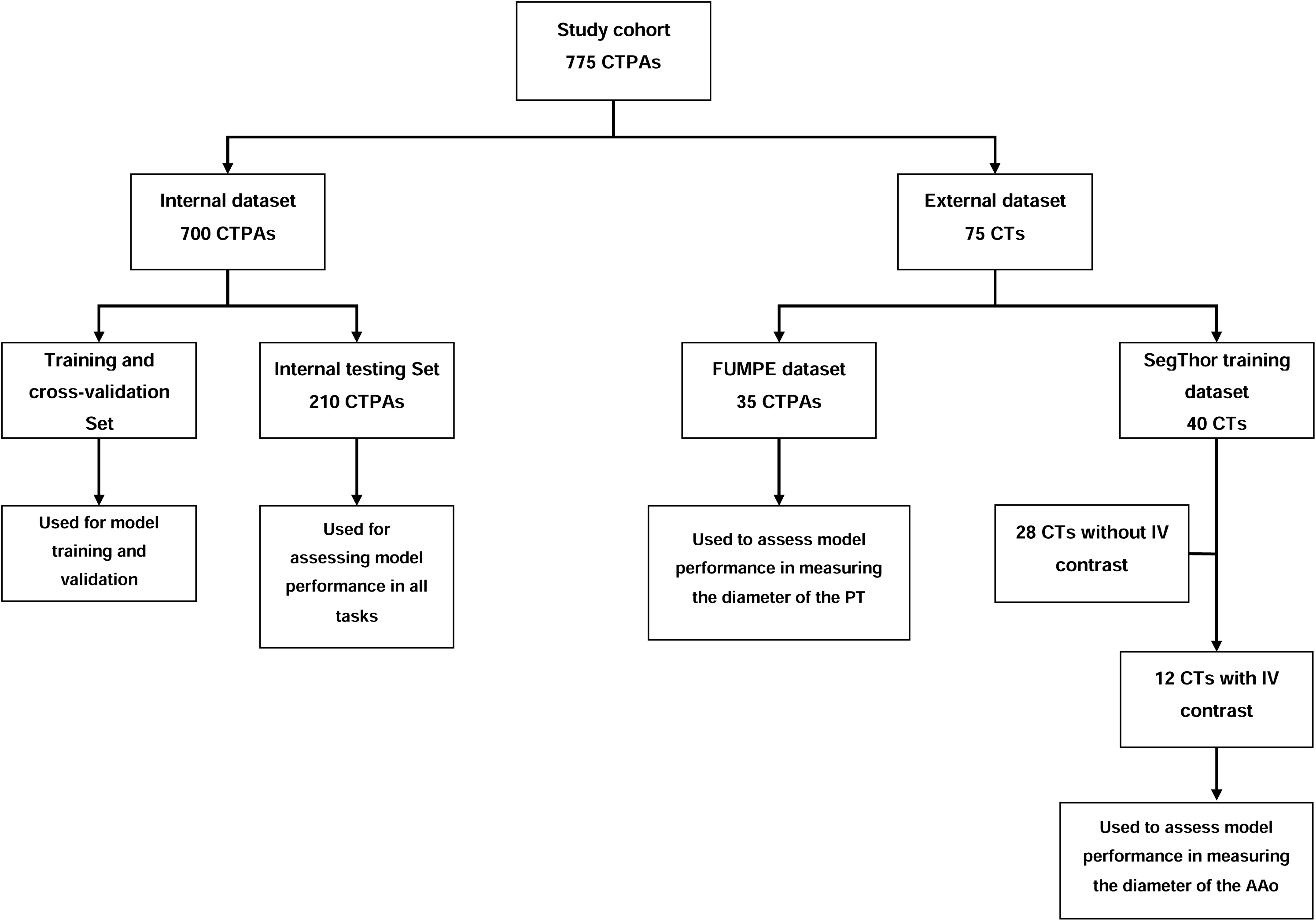
Schematic of the study cohort. From the internal dataset, 210 CT pulmonary angiography (CTPA) examinations were used for model testing across all tasks: measurement of image noise, intravenous (IV) contrast in the pulmonary trunk (PT), diameter of the ascending aorta (AAo), and diameter of the PT. The FUMPE dataset was used for external validation of PT diameter measurements. From the SegThor dataset, 28 CT examinations without IV contrast were excluded. The remaining 12 CT scans with IV contrast were used to test the diameter of the AAo.

### Computational Environment

The computational environment used for both model training and inference on test data in this study includes specific software versions: Ubuntu (22.04.3 LTS) as the operating system, Docker (27.0.3) for containerization, and CUDA (12.1.1), along with Nvidia driver (535.183.01), to ensure seamless interaction with NVIDIA GeForce RTX 2080 Ti GPUs. The programming languages used are Python (3.10.6) and MATLAB R2023b (The MathWorks, Inc., Natick, Massachusetts, USA). The deep learning framework PyTorch (version 2.1.0) was utilized, and the nnU-Net framework was implemented in version 2.1.1. Post-processing computations were conducted on the same machine equipped with an Intel Core i9-9900X CPU (10 cores, 20 threads, 3.5 GHz base clock, 4.5 GHz boost clock).

### Model training and validation

For model training, the nnU-Net framework was implemented using the PyTorch Nvidia container (nvcr.io/nvidia/pytorch:24.04-py3) on a Docker platform (Docker Inc., Palo Alto, California, USA), providing an optimized environment for deep learning and scientific computing. The nnU-Net deep learning open-source framework is an adaptable and automated solution designed for semantic segmentation tasks, particularly in the field of medical image segmentation. It builds on a U-Net-like architecture with a contracting path (encoder) and an expansive path (decoder) with skip connections to efficiently capture and process image details. The framework automates the processes of data preprocessing, training pipeline, and network configuration, including aspects such as network topology, depth, learning rate, filter selection, patch and batch size, based on the dataset characteristics, thereby eliminating the need for manual fine-tuning [18].

A 2D U-Net architecture with a depth of 7 was generated by the nnU-Net framework, designed for an initial input size of 512×512 pixels (Figure 2). In the downsampling operations (contracting path), each level consisted of a 2D convolutional layer with a kernel size of 3×3 and a stride of 2, followed by another 2D convolutional layer with the same kernel size but a stride of 1. The network started with 32 filters and increased up to 512 filters as the depth increases. In the upsampling operations (expansive path), a 2D transposed convolutional layer with a kernel size of 2×2 and a stride of 2 was used. Both downsampling and upsampling operations were followed by instance normalization and a leaky rectified linear unit (Leaky ReLU) activation.

**Figure 2.**
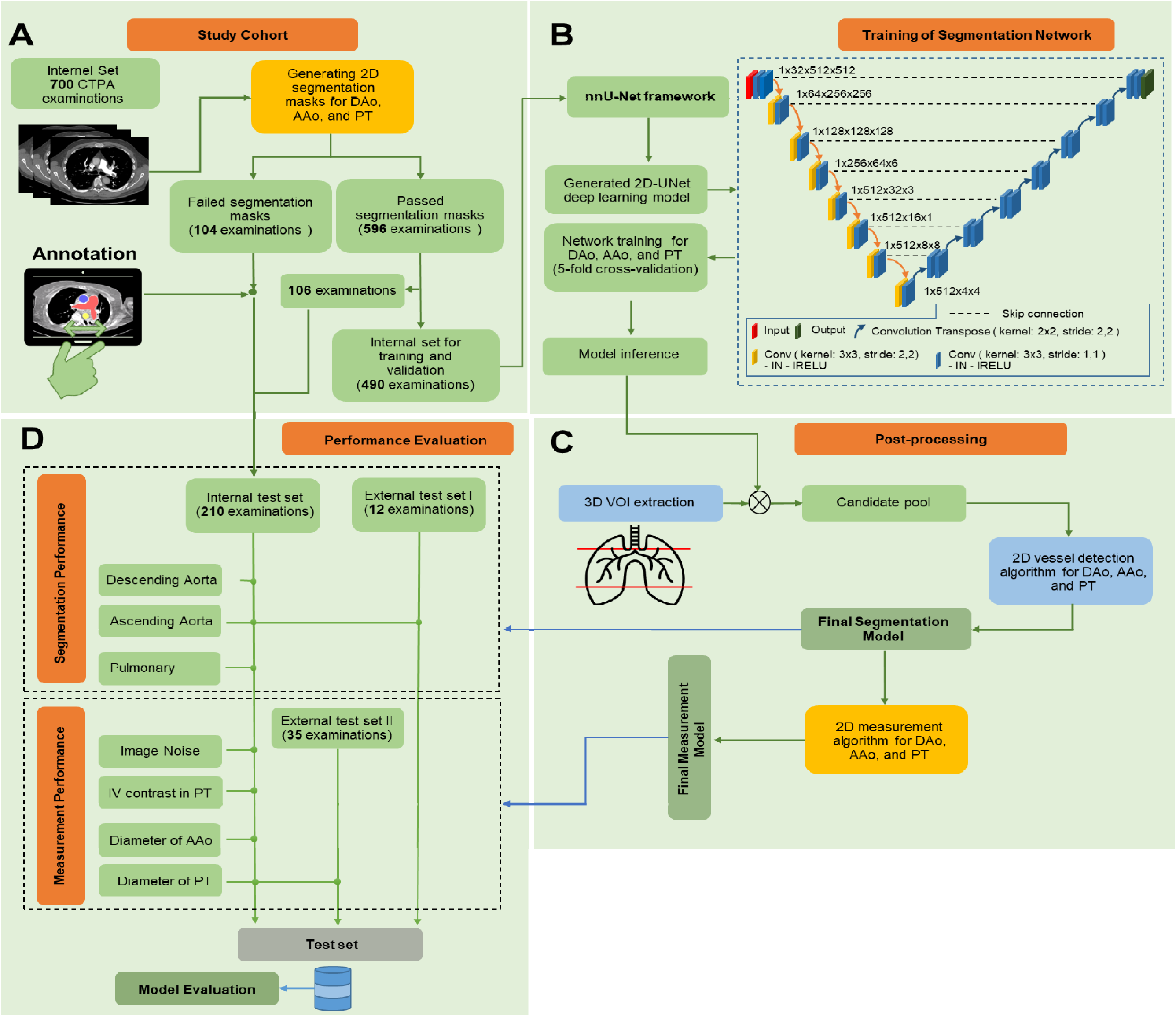
Study Design. 700 CT pulmonary angiography (CTPA) examinations were collected, and a fully automated deterministic segmentation algorithm [19] was applied to generate 2D segmentation masks for the descending aorta (DAo), ascending aorta (AAo), and pulmonary trunk (PT). Segmentation masks were successfully generated for 596 of these examinations, while the remaining 104 required manual annotation. The dataset was divided into a training set comprising 490 examinations and a testing set comprising 210 examinations (a). The 2D U-Net deep learning model, generated by the nnU-Net framework, was trained on 490 CTPA scans using 5-fold cross-validation. By default, the convolution layer used a 3×3 filter size, followed by an instance normalization (IN) layer and a leaky rectified linear unit (lRELU) layer (b). Model inference was applied to the volume of interest (VOI) to generate a pool of vessel candidates. A 2D vessel detection algorithm was then applied to the candidate pool to determine the objects of interest for measurement. A 2D measurement algorithm [19] was subsequently applied to achieve accurate vessel measurements (c). For segmentation and measurement performance, the final model was evaluated on 210 CTPAs from the internal dataset and on 12 and 35 CTPAs from two publicly available datasets, respectively (d). Modules in gold-colored boxes represent previously developed solutions, while those in light blue-colored boxes were developed as part of this work.

During training, the nnU-Net framework utilized a tailored set of hyperparameters to optimize the learning process. The optimizer selected was Stochastic Gradient Descent (SGD) with Nesterov momentum, with a momentum value of 0.99. Weight decay was applied with a coefficient of 3e-05 to control model complexity. The training process was set to run for a maximum of 1000 epochs. To further refine the training, a learning rate scheduler was used, which progressively reduced the learning rate each epoch, starting at 0.01 and decreasing to 2e-05 by the final epoch. The nnU-Net framework employs a variety of data augmentation techniques to enhance model generalization and prevent overfitting to the training dataset. These techniques include spatial transformations such as rotations, scaling, and mirroring. Specifically, the rotations cover a full 360-degree range around the X-axis, with scaling applied between 0.7 to 1.4 times the original size. Mirroring is applied along both the X and Y axes. Additionally, Gaussian noise and blur, as well as brightness, contrast, and gamma transformations, are utilized to further augment the data and improve model robustness.

### Model inference and post-processing

After model training, three trained models were obtained for the segmentation tasks of the DAo, AAo, and PT. Before applying model inference, the volume of interest (VOI) was first determined by including only the slices between approximately 22% and 65% of the total size of the CTPA volume. This approach ensured that most of the mediastinum was accurately captured, encompassing two key anatomical landmarks: the carina of the trachea and the level of the pulmonary trunk. Model inference was then applied only to this volume of interest to generate candidate pools for DAo, AAo, and PT segmentations. Once the candidates were obtained, deterministic algorithms were developed in MATLAB to identify the best 2D segmentation masks for measurement. The study design is illustrated in Figure 2.

### Descending aorta segmentation

To ensure the correct selection of the descending aorta (DAo) from the segmentation candidates, we first applied 3D connected component analysis to identify the largest component within the volume of interest (VOI), assuming it to be the DAo. Next, we removed any connected components with fewer than 200 pixels using 8-connectivity and performed a morphological closing operation on the binary image with a disk-shaped structuring element having a radius of 9 pixels. These image processing steps focused on isolating and refining the largest 3D structure, presumably the DAo, by filtering out smaller components and smoothing the object boundaries. Once we obtained the 3D segmentation of the DAo, we applied the same measurement algorithm used by radiologists. Specifically, the standard deviation of HU within a 1 cm² circular region of interest (ROI) in the DAo was used as a measure of image noise in the CTPA examination. We calculated the image noise in every 2D segment of the 3D DAo segmentation and then took the median of these values. The median value of the image noise provided the final measurement.

### Ascending Aorta Segmentation

We began by applying a filtering operation to eliminate small, irrelevant objects from the candidate pool using an area opening operation, which removes connected components with fewer than 200 pixels based on 8-connectivity. Next, we performed a 3D connected component analysis, retaining only components with an area greater than 12,500 pixels. To ensure the spatial relevance of the selected component, likely representing the AAo, we calculated the z-coordinate of the centroids of the filtered components. The component whose centroid was closest to the superior part of the CTPA was identified and retained for further processing. We then tracked any sudden area loss or significant shifts in centroid position to remove adjacent objects or irrelevant tissues. After these operations, a final connected component analysis using 26-connectivity was performed to select the largest remaining component in the candidate pool. Finally, a morphological closing operation with a disk-shaped structuring element with a radius of 9 pixels was applied to smooth the boundaries of the final 3D component. To measure the diameter of the AAo, we used the equivalent diameter property of the ‘regionprops’ function in MATLAB’s image processing toolbox. We calculated the equivalent diameter for all 2D objects within the final 3D component, and the median value of these 2D equivalent diameters provided the final measurement.

### Pulmonary Trunk (PT) Segmentation and IV contrast in PT

We first eliminated components smaller than 200 pixels from the candidate pool. The candidate pool was further refined by applying morphological opening using a disk-shaped structuring element with a radius of 7 pixels, helping to remove noise and small irrelevant structures. Next, components with less than 10,000 pixels were removed, ensuring that only significant regions remain. To identify the components most likely corresponding to the pulmonary trunk within the candidate pool based on the slice number of the CTPAs, we established two reference points as follows:

Let *S* represent the total number of slices in the CTPA. Then:

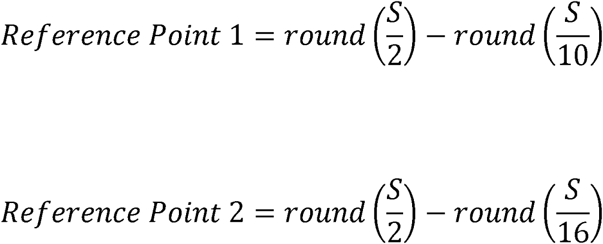

The algorithm then calculates the distance of each component’s centroid from the reference points and counts the number of slices above, below, and within the reference regions. If exactly two components are present, the algorithm evaluates their proximity to the reference points and, based on the patient’s position, selects the component that is either the highest or lowest in the scan volume as the pulmonary trunk. For three components, the algorithm prioritizes them according to their proximity to the reference points and selects the most appropriate one based on the patient’s position. If more than three components are identified, the algorithm refines its selection criteria further, focusing on the second or third closest component, depending on the patient’s position, to accurately identify the pulmonary trunk. Finally, the selected component undergoes a closing operation using a disk-shaped structuring element with a radius of 9 pixels to smooth the boundaries and complete the segmentation of the pulmonary trunk. As a measure of the IV contrast concentration in the PT, we calculated the mean HU of all 2D objects within the final 3D component. To measure the diameter of the PT, we extracted horizontal line segments based on the Hough transform for each 2D object within the final 3D component, and the median value of these 2D Hough lines provided the final diameter measurement.

### Statistical Analysis

The model’s segmentation performance was evaluated using the Boundary F1 (BF) score, the Dice-Sørensen coefficient (DSC), and the Jaccard Index (JI) with Matlab (MathWorks, Inc., R2023b). The BF score evaluates the accuracy of the model segmentation boundary of an object by comparing it to the ground truth segmentation boundary [23]. The BF score is calculated as the harmonic mean of recall and precision:

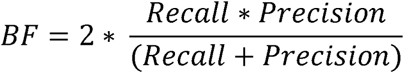

Here, *Recallll* is the ratio of ground truth boundary points near the model segmentation boundary to the total ground truth segmentation boundary length. *Precision* is the ratio of model boundary points near the ground truth segmentation boundary to the total model segmentation boundary length.

Given two sets, A (model segmentation) and B (ground truth segmentation),

DSC is defined as:

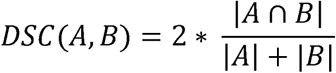

And JI defined as follow:

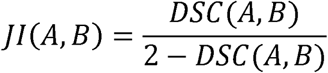

Bland-Altman [24] and scatter plot analyses were used to evaluate measurement performance of the model. Specifically, Bland-Altman analysis assessed the agreement between the model’s and the radiologist’s measurements, while the scatter plot analysis examined the relationship between them. Pearson’s correlation coefficient was utilized to evaluate the correlation between the model’s and the radiologist’s measurements. Subgroup analysis of radiologist measurements, based on examination quality, sex, and age, was conducted using the Kruskal-Wallis H-test. A p-value of less than 0.05 was considered statistically significant. For statistical analysis, we utilized SciPy, a Python library for scientific computing (version 1.13.1), and Microsoft Excel from Microsoft 365 (Microsoft Corporation, Washington, USA).

## Results

The internal dataset (n=700 CTPAs) included 383 CTPA examinations from 353 women (median age, 73 years; IQR, 20 years) and 317 examinations from 299 men (median age, 71 years; IQR 15 years) (**Table 1**). The internal dataset was split into training and testing sets using a 70/30 split ratio. For the training and 5-fold cross-validation sets, 1,470 2D segmentation masks from 490 CTPAs were selected. The remaining 630 2D segmentation masks from 210 CTPAs were used for the testing set. Additionally, all 312 manually annotated 2D segmentation masks from 104 CTPAs were included in the testing set. There is no patient overlap between the training and testing sets. For model training, each of the three major blood vessels (DAo, AAo, and PT) was trained separately using the internal dataset comprising 1,470 2D masks (490 masks per vessel) derived from 490 CTPAs. The segmentation models were trained using 5-fold cross-validation, with 80% of the data (a total of 1,176 2D masks, 392 masks per vessel) used for training and 20% (a total of 294 2D masks, 98 masks per vessel) used for cross-validation.

**Table 1.** Study Characteristics of the Internal Dataset.

| Characteristics | Training Dataset | Test Dataset | Overall |
| --- | --- | --- | --- |
| No. of patients | 466 | 186 | 652 |
| No. of CTPA exams | 490 | 210 | 700 |
| Sex (Women/Men) | 245/221 | 108/78 | 353/299 |
| Median age (IQR) years | 72.0 (17.0) | 74.0 (19.75) | 72.0 (18.0) |
| Women | 73.0 (19.0) | 74.0 (22.5) | 73.0 (20.0) |
| Men | 71.0 (16.0) | 74.0 (16.0) | 71.0 (15.0) |
| Diseases |  |  |  |
| Emboli in (Right/Left) lung | 100/81 | 37/33 | 137/114 |
| Pneumothorax in (Right/Left) lung | 4/2 | 0/0 | 4/2 |
| Pleural effusion in (Right/Left) lung | 147/140 | 83/74 | 230/214 |
| Infiltrate or atelectasis (or other opacities) in (Right/Left) lung | 298/275 | 146/138 | 444/413 |
| Pericardial fluid (excessive amounts) | 57 | 26 | 83 |
| Sign of acute congestive heart failure | 89 | 42 | 131 |
| Lymph nodes (Mediastinum/Right hilum/Left hilum)<sup>1</sup> | 90/48/37 | 48/28/17 | 138/76/54 |
| Quality of CTPA exams (Good/Acceptable/Inferior) | 200/158/132 | 58/70/82 | 258/228/214 |

Note. — Unless otherwise indicated, data are number of examinations. CTPA = computed tomography pulmonary angiography
<sup>1</sup> Lymph nodes larger than 1 cm (short axis)

### Radiologists Ground Truth Measurements

The median image noise was 20.5 HU (IQR, 8 HU) in the internal training dataset and 20 HU (IQR, 10 HU) in the internal test dataset, with significant differences observed based on examination quality, sex, and age groups older than 64 years (**Table 2 and Supp. Table 1**). The median diameter of the AAo was 33 mm (IQR, 6 mm) in the internal training dataset and 34 mm (IQR, 6 mm) in the internal test dataset, with significant differences particularly noted across sex and older age groups. The median IV contrast concentration in PT was 394 HU (IQR, 146 HU) in the internal training dataset and 358 HU (IQR, 158 HU) in the internal test dataset, with significant differences observed in older patients, especially those aged over 79 years. The median diameter of the PT was 26 mm (IQR, 5 mm) in the internal training dataset and 28 mm (IQR, 6 mm) in the internal test dataset, without significant differences across examination quality, sex, or age groups.

**Table 2.** Radiologists Ground Truth Measurements.

| Category | Training Dataset (n=490) |  |  |  |  | Test Dataset (n=210) |  |  |  |  |
| --- | --- | --- | --- | --- | --- | --- | --- | --- | --- | --- |
|  | Quality of CTPA exams |  |  |  | Overall | Quality of CTPA exams |  |  |  | Overall |
|  | Good | Acceptable | Inferior | p-value |  | Good | Acceptable | Inferior | p-value |  |
| No. of CTPA exams <sup>1</sup> |  |  |  |  |  |  |  |  |  |  |
| Women | 121 (24.69) | 80 (16.33) | 56 (11.43) |  | 257 (52.45) | 41 (19.52) | 41 (19.52) | 44 (20.95) |  | 126 (59.99) |
| Men | 79 (16.12) | 78 (15.92) | 76 (15.51) |  | 233 (47.55) | 17 (8.1) | 29 (13.81) | 38 (18.1) |  | 84 (40.01) |
| Age < 65 years | 62 (12.65) | 50 (10.2) | 33 (6.73) | - | 145 (29.58) | 16 (7.62) | 21 (10) | 23 (10.95) | - | 60 (28.57) |
| Age 65-79 years | 92 (18.78) | 78 (15.92) | 61 (12.45) |  | 231 (47.15) | 25 (11.9) | 31 (14.76) | 30 (14.29) |  | 86 (40.95) |
| Age ≥ 80 years | 46 (9.39) | 30 (6.12) | 38 (7.76) |  | 114 (23.27) | 17 (8.1) | 18 (8.57) | 29 (13.81) |  | 64 (30.48) |
| Image noise (HU) <sup>2</sup> |  |  |  |  |  |  |  |  |  |  |
| Overall | 19.0 (9.0) | 20.0 (7.0) | 23.0 (7.0) | < 0.0001 | 20.5 (8.0) | 16.0 (9.8) | 20.0 (8.8) | 23.5 (8.0) | < 0.0001 | 20.0 (10.0) |
| Women | 19.0 (10.0) | 20.5 (7.0) | 21.0 (8.0) | 0.02 | 20.0 (9.0) | 16.0 (9.0) | 20.0 (7.0) | 23.0 (8.0) | 0.0004 | 20.0 (10.0) |
| Men | 20.0 (7.0) | 20.0 (7.0) | 23.5 (5.2) | 0.0002 | 21.0 (8.0) | 16.0 (9.0) | 19.0 (10.0) | 24.0 (8.8) | 0.003 | 21.0 (9.5) |
| Age < 65 years | 21.0 (8.0) | 19.0 (7.8) | 21.0 (8.0) | 0.92 | 20.0 (8.0) | 15.5 (12.2) | 18.0 (8.0) | 22.0 (6.5) | 0.11 | 20.0 (9.5) |
| Age 65-79 years | 18.5 (9.2) | 20.5 (7.8) | 23.0 (6.0) | 0.0001 | 20.0 (9.0) | 18.0 (7.0) | 20.0 (8.0) | 24.0 (7.8) | 0.006 | 20.0 (8.8) |
| Age ≥ 80 years | 19.0 (9.8) | 20.5 (6.5) | 23.5 (4.8) | 0.001 | 21.0 (7.8) | 14.0 (5.0) | 21.0 (9.5) | 24.0 (9.0) | 0.0005 | 20.0 (10.8) |
| Diameter of ascending aorta (mm) <sup>3</sup> |  |  |  |  |  |  |  |  |  |  |
| Overall | 32.0 (6.0) | 33.0 (5.0) | 34.0 (6.0) | 0.01 | 33.0 (6.0) | 35.0 (6.5) | 33.0 (6.0) | 34.0 (6.0) | 0.07 | 34.0 (6.0) |
| Women | 31.0 (5.0) | 32.0 (5.0) | 32.0 (7.0) | 0.45 | 31.0 (6.0) | 34.0 (6.0) | 31.0 (7.0) | 32.0 (4.0) | 0.11 | 33.0 (6.5) |
| Men | 34.0 (6.0) | 35.0 (4.8) | 35.0 (5.2) | 0.22 | 35.0 (5.0) | 38.0 (7.0) | 35.0 (4.0) | 36.0 (4.8) | 0.07 | 36.0 (5.0) |
| Age < 65 years | 29.5 (6.0) | 31.0 (5.8) | 31.0 (8.0) | 0.3 | 31.0 (7.0) | 32.0 (7.8) | 29.0 (6.0) | 31.0 (5.0) | 0.39 | 31.0 (7.0) |
| Age 65-79 years | 34.0 (5.2) | 34.0 (4.0) | 34.0 (4.0) | 0.72 | 34.0 (5.0) | 35.0 (7.0) | 33.0 (6.5) | 35.0 (4.0) | 0.28 | 35.0 (6.0) |
| Age ≥ 80 years | 33.0 (6.0) | 35.0 (5.8) | 36.0 (5.8) | 0.01 | 35.0 (7.0) | 37.0 (3.0) | 35.5 (3.5) | 36.0 (6.0) | 0.34 | 36.0 (5.0) |
| IV contrast concentration in PT (HU) □ |  |  |  |  |  |  |  |  |  |  |
| Overall | 425 (125) | 370 (134) | 374 (193) | < 0.0001 | 394 (146) | 406 (141) | 334 (148) | 316 (167) | < 0.0001 | 358 (158) |
| Women | 432 (123) | 363 (135) | 351 (189) | < 0.0001 | 390 (144) | 404 (98) | 343 (122) | 372 (212) | 0.03 | 372 (145) |
| Men | 419 (128) | 380 (124) | 398 (194) | 0.03 | 396 (143) | 493 (156) | 321 (158) | 296 (138) | 0.003 | 322 (172) |
| Age < 65 years | 406 (127) | 326 (102) | 280 (81) | < 0.0001 | 356 (129) | 372 (76) | 321 (90) | 294 (167) | 0.03 | 326 (132) |
| Age 65-79 years | 422 (103) | 387 (141) | 386 (172) | 0.08 | 396 (136) | 435 (148) | 335 (140) | 312 (132) | 0.004 | 356 (150) |
| Age ≥ 80 years | 448 (121) | 394 (120) | 454 (136) | 0.05 | 441 (140) | 482 (156) | 392 (166) | 398 (227) | 0.22 | 406 (224) |
| Diameter of PT (mm) <sup>3</sup> |  |  |  |  |  |  |  |  |  |  |
| Overall | 26.0 (4.0) | 27.0 (6.0) | 27.0 (5.0) | 0.002 | 26.0 (5.0) | 27.0 (7.0) | 27.5 (5.0) | 29.0 (6.0) | 0.05 | 28.0 (6.0) |
| Women | 25.0 (5.0) | 26.5 (5.0) | 27.0 (4.2) | 0.02 | 26.0 (6.0) | 26.0 (6.0) | 27.0 (5.0) | 28.0 (4.5) | 0.24 | 27.0 (5.8) |
| Men | 26.0 (5.5) | 28.0 (5.0) | 27.0 (6.0) | 0.33 | 27.0 (5.0) | 29.0 (3.0) | 29.0 (8.0) | 30.0 (4.8) | 0.45 | 29.0 (5.2) |
| Age < 65 years | 25.0 (4.8) | 26.0 (6.0) | 27.0 (5.0) | 0.11 | 26.0 (5.0) | 25.5 (6.2) | 25.0 (6.0) | 28.0 (7.5) | 0.06 | 26.0 (5.2) |
| Age 65-79 years | 26.0 (5.0) | 27.0 (5.0) | 27.0 (5.0) | 0.24 | 27.0 (5.0) | 28.0 (8.0) | 27.0 (4.0) | 29.0 (4.8) | 0.2 | 28.0 (6.0) |
| Age ≥ 80 years | 26.0 (4.0) | 28.0 (8.5) | 28.0 (6.8) | 0.04 | 27.0 (6.0) | 29.0 (5.0) | 30.5 (5.2) | 29.0 (5.0) | 0.43 | 29.0 (5.2) |

Note. — Unless otherwise indicated, data are number of examinations and data in parentheses are the interquartile range., HU = hounsfield unit, mm = millimeter, PT = pulmonary trunk, IV = intravenous, ROI= region of interest
<sup>1</sup> Data in parentheses are the percentage.
<sup>2</sup> The measurement was done by calculating the SD of HU within a circular ROI of 1 cm<sup>2</sup> in the descending aorta at the level of the PT.
<sup>3</sup> The diameters of the PT and ascending aorta were measured at the level of the PT.
□ The measurement was done by calculating the mean HU in a circular ROI of 2 cm<sup>2</sup> just proximal to the PT bifurcation.

### Model segmentation performance evaluation on the internal and external testing dataset

Segmentation performance was evaluated for three major blood vessels: the descending aorta (DAo), ascending aorta (AAo), and pulmonary trunk (PT), on an internal test dataset of 210 CTPAs. The trained nnU-Net deep learning model successfully detected all three major blood vessels in all 210 CTPAs. However, there were segmentation failures in 3 cases for the DAo, 8 cases for the AAo, and 15 cases for the PT. For the successfully segmented vessels, the median Dice scores for the DAo were 0.94 (IQR, 0.05) for good quality, 0.95 (IQR, 0.05) for acceptable quality, and 0.95 (IQR, 0.04) for inferior quality examinations. For the AAo, the scores were 0.96 (IQR, 0.04), 0.96 (IQR, 0.05), and 0.95 (IQR, 0.05), respectively. Similarly, for the PT, the scores were 0.96 (IQR, 0.05), 0.95 (IQR, 0.04), and 0.95 (IQR, 0.04), corresponding to good, acceptable, and inferior quality examinations. A combined overall median Dice score of 0.95 (IQR, 0.05) for these three major blood vessels was achieved on the internal test dataset (**Figure 3, A**). AAo segmentation was performed on 12 contrast-enhanced CT examinations from the SegTHOR dataset [21]. The median Dice, Jaccard, and Boundary F1 (BF) contour matching scores were 0.94 (IQR, 0.02), 0.88 (IQR, 0.04), and 1.0, respectively (**Figure 3, B)**.

**Figure 3.**
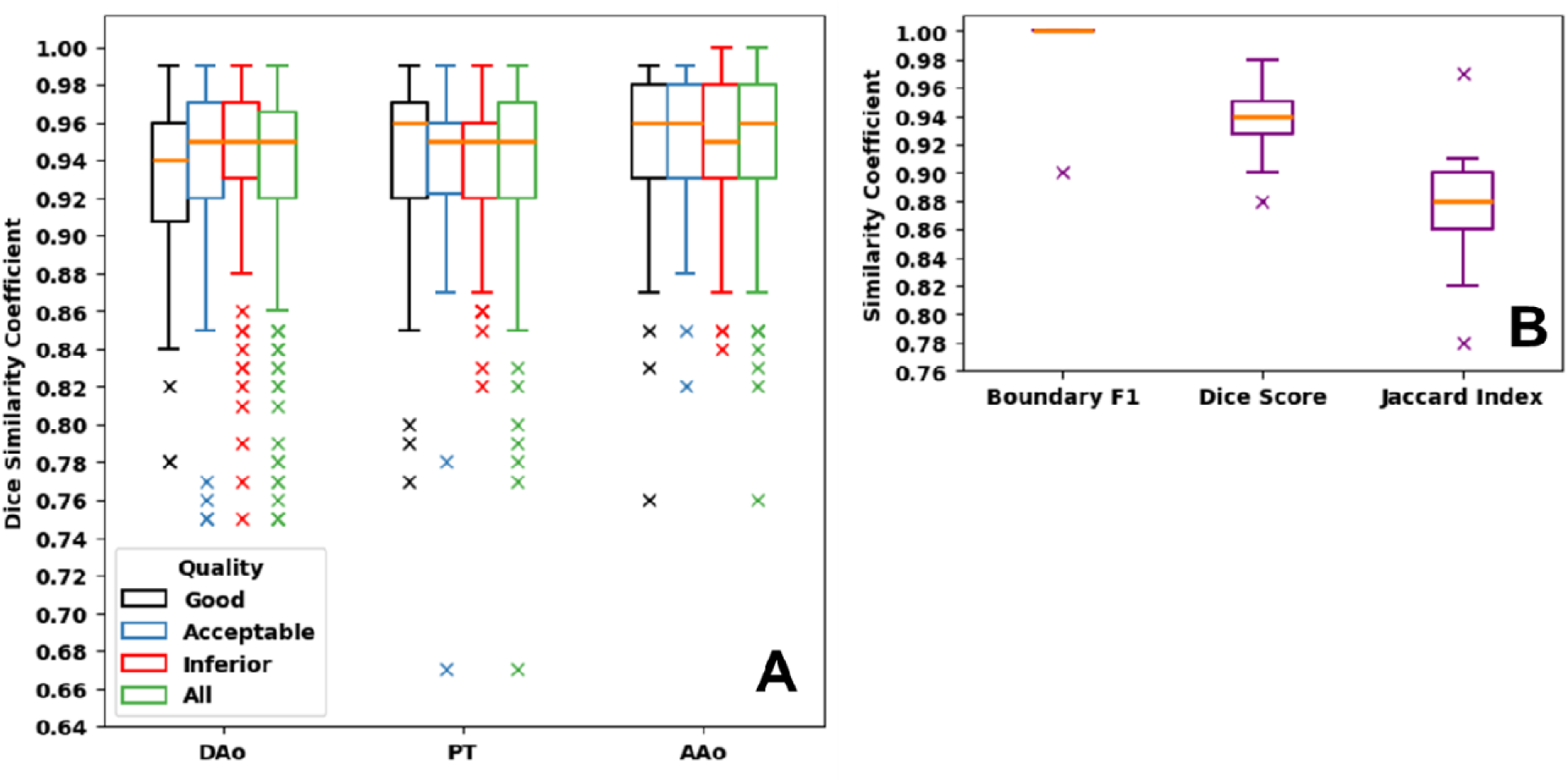
Segmentation performance on the internal and external test sets. (A) The internal test set segmentation performance, measured by the Dice Similarity Coefficient (DSC), is provided for all tasks, including the descending aorta (DAo, n= 207 CTPA exams), the pulmonary trunk (PT, n= 195 CTPA exams), and the ascending aorta (AAo, n= 202 CTPA exams). The quality of each CTPA examination was rated by a radiologist as good (black), acceptable (blue), or inferior (red). Green boxes represent the DSC for all quality ratings. (B) The Boundary F1, Dice, and Jaccard index scores for AAo segmentation are provided for the SegTHOR dataset (nℒ=ℒ12 CT exams with contrast agent). The median (orange line), interquartile range (boxes), and outliers (x) are shown.

### Model measurement performance evaluation on the internal and external testing dataset

Measurement evaluations were conducted on the successfully segmented vessels. Image noise was assessed on 207 CTPAs, the diameter of the AAo on 202 CTPAs, and both the IV contrast concentration in PT and the diameter of the PT on 195 CTPAs from the internal testing dataset. These measurements resulted in Pearson’s r values of 0.91, 0.93, 0.98, and 0.55, respectively, all with p < 0.001 (**Figure 4, A1-D1)**. The proposed model’s measurements compared favorably with those of the radiologist, with mean differences observed for image noise (−0.03 HU), AAo diameter (1.78 mm), PT IV contrast level (17.62 HU), and PT diameter (−2.16 mm) (**Figure 4, A2-D2).** We also present the mean percentage differences between the proposed model and the radiologist using Bland-Altman analysis: −1.96% for image noise, 5.56% for the AAo diameter, and −7.65% for the PT diameter) (**Figure 4, A3-D2)**.

**Figure 4.**
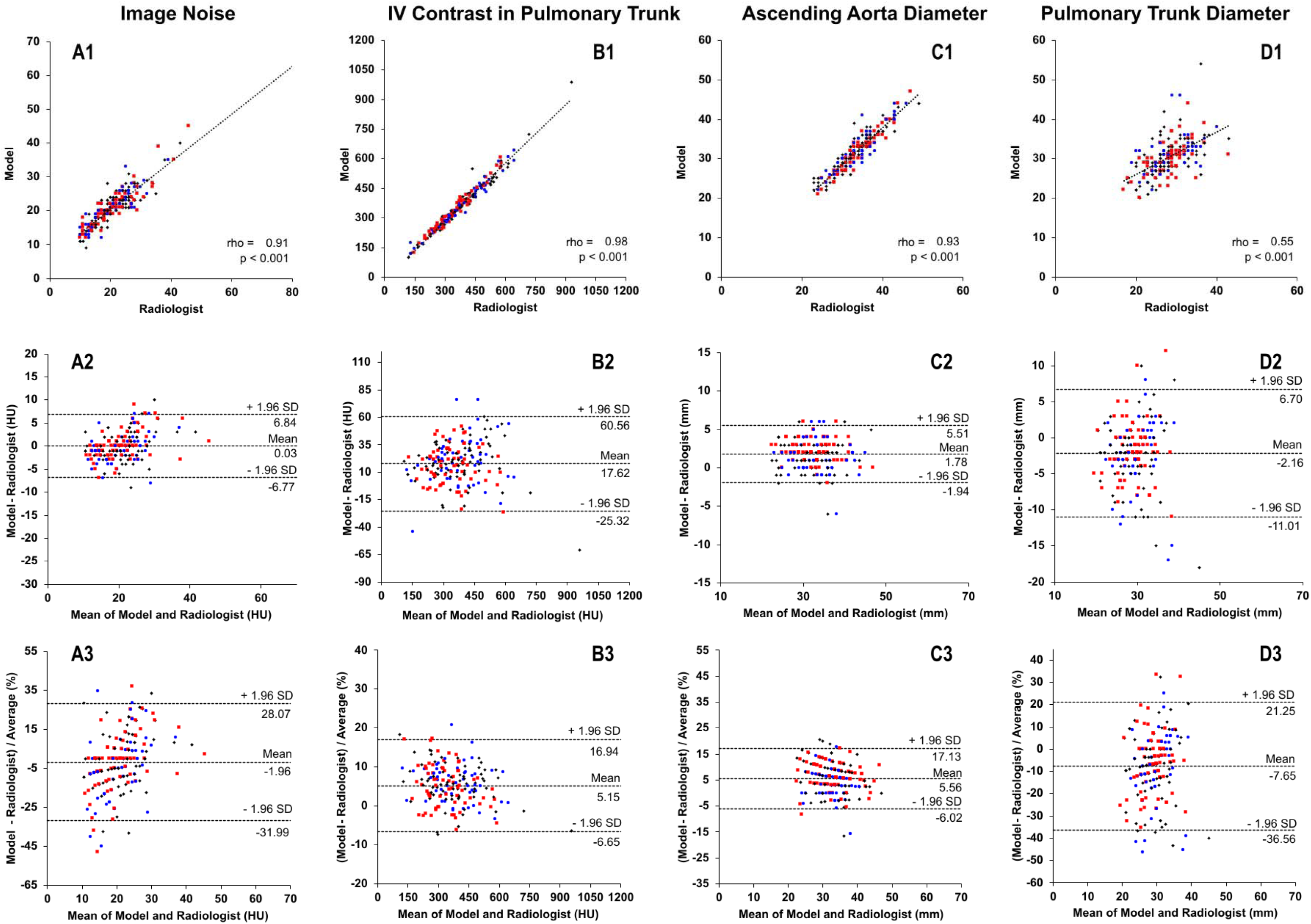
Comparison of developed AI model to the Radiologist. Both automatic and manual measurements from the test set cases were evaluated using regression analysis (top row, dashed regression lines), Bland-Altman plots for differences in radiodensity and diameters (middle row, limits of agreement from −1.96 to +1.96 SD), and Bland-Altman plots for percentage differences (bottom row, limits of agreement from −1.96 to +1.96 SD). The analysis included: (A) Image noise (n=207 CTPA exams), (B) Intravenous (IV) contrast agent in pulmonary trunk (n=195 CTPA exams), (C) Ascending aorta diameter (n=202 CTPA exams), and (D) Pulmonary trunk diameter (n=195 CTPA exams). The quality of each CTPA examination was rated by the radiologist as good (black diamonds), acceptable (blue circles), or inferior (red squares).

For the external evaluation of measurements, PT diameter was assessed in 35 CTPA exams from the FUMPE dataset. First, inference was performed using a trained nnU-Net DL model, followed by the selection of the best 2D PT segmentation candidate through a post-processing algorithm. Finally, a measurement algorithm was applied, resulting in Pearson’s r = 0.83, p < 0.001 (**Figure 5, A)**. The limits of agreement between the proposed model and FUMPE radiologist annotations showed mean differences of −1.43 mm and −5.14% (**Figure 5, B and C).**

**Figure 5.**
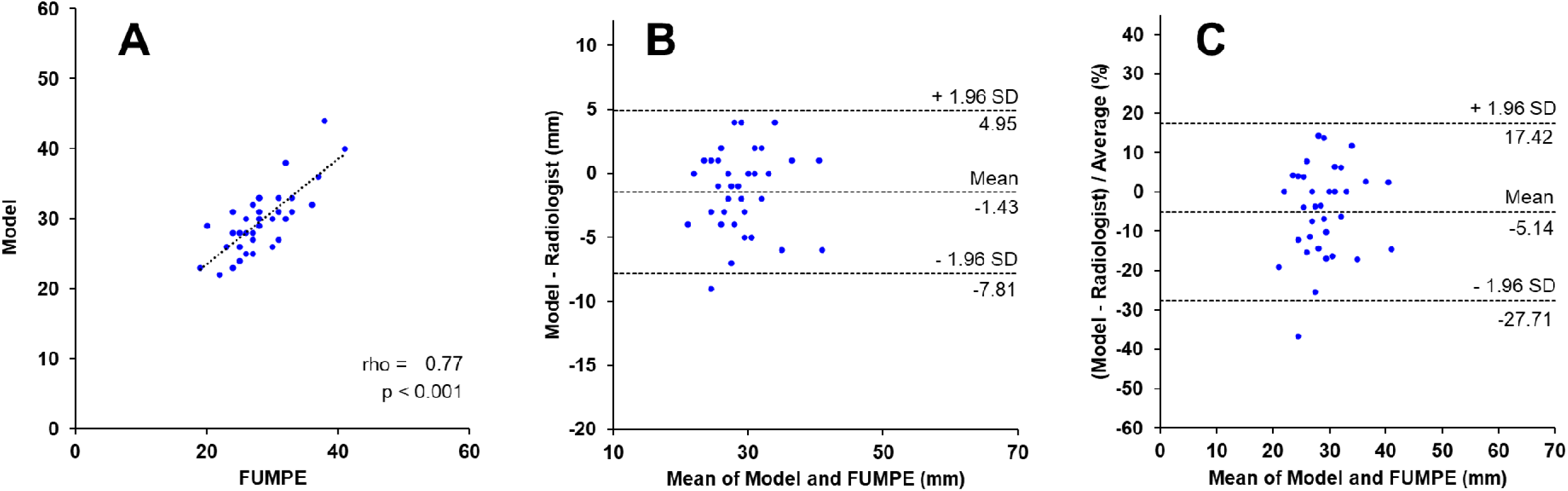
External Testing of Automatic Pulmonary Trunk (PT) Diameter Measurement. Automatic PT diameter measurements from the FUMPE dataset (35 CTPA exams) were compared to manual radiologist annotations from the original study using three methods: (A) regression analysis, depicted with dashed regression lines, (B) Bland-Altman plots of differences in millimeters, with limits of agreement from −1.96 to +1.96 SD, and (C) Bland-Altman plots of percentage differences, with the same limits of agreement.

### Segmentation and measurement performance comparison with the previous solution

This study demonstrates significant improvements in segmentation performance over our previously proposed fully automated deterministic solution [19], particularly with higher success rates for segmenting the DAo (99% vs. 90%), AAo (96% vs. 86%), and PT (93% vs. 88%) in the internal test set. Measurement performance had notable improvements in image noise (0.03 HU vs. −0.25 HU) and the diameter of the PT (−2.16 mm vs. −3.20 mm), but a larger variance in the diameter of the AAo (1.78 mm vs. 0.51 mm) and IV contrast concentration in the PT (17.62 HU vs. −0.28 HU) (**Table 3**). Overall, the current study provides enhanced segmentation accuracy, while measurement accuracy improvements are more variable.

**Table 3.**
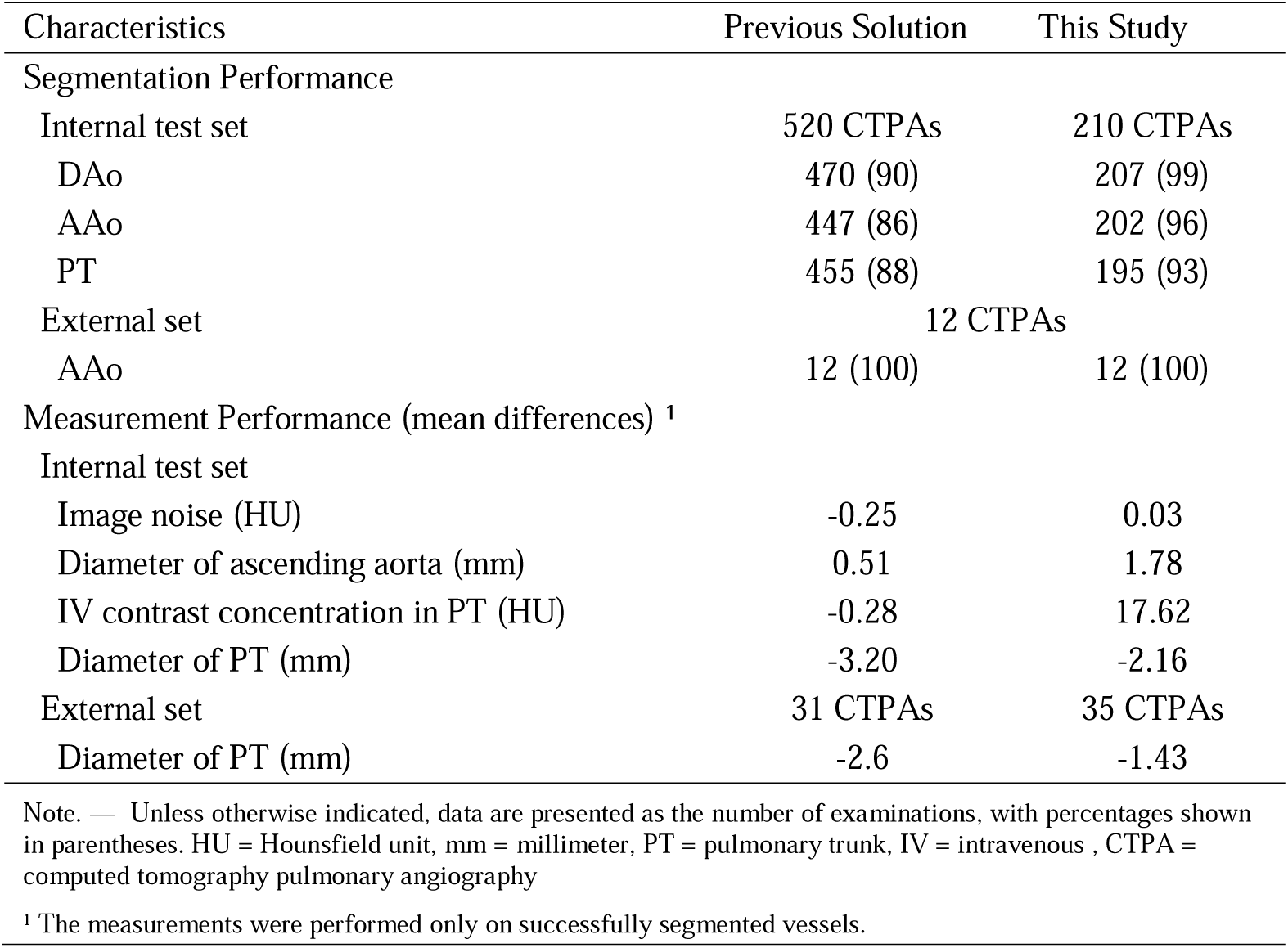
Comparison of Segmentation and Measurement Performance Between the Current and Previous Solution.

## Discussion

In this study, a nnU-Net [18] based deep learning (DL) model was used along with a deterministic post-processing step to automatically segment major blood vessels including descending aorta (DAo), ascending aorta (AAo), and pulmonary trunk (PT). The proposed model achieved robust segmentation of major blood vessels, with a combined overall median Dice score of 0.95 (interquartile range, 0.05). For successfully segmented vessels, a 2D measurement algorithm [19] was adapted and subsequently applied to achieve accurate vessel measurements. The automatic measurements showed a strong correlation with the radiologist’s assessments, with Pearson correlation coefficients ranging from 0.55 to 0.98.

The comparison of segmentation methods for vascular structures, including traditional image processing approaches such as multi-atlas segmentation and model-based frameworks as well as modern artificial intelligence methods like deep learning models, revealed varying performance across different techniques. Our proposed method significantly outperformed our previous fully automated deterministic solution [19], particularly in segmenting the DAo, AAo, and PT. Segmentation success rates increased by 9%, 10%, and 5%, respectively, on the internal test set. Zhuang et al. employed multi-atlas segmentation on 30 cases, achieving high dice score for the AAo (0.96) but lower performance for the pulmonary trunk (0.79), highlighting the challenge of segmenting complex structures. In contrast, Ecabert et al. used a model-based framework with geometric mesh modeling and deformable models on 37 cases, delivering strong and consistent scores for both the aorta (0.95) and PT (0.94), indicating the robustness of their method in 3D segmentation tasks. Deep learning approaches, particularly U-Net-based semantic segmentation architectures, have demonstrated outstanding performance in numerous studies [15,25–27]. Baskaran et al. [15] tested their U-Net model on 20 cases, achieving high Dice scores for the aorta (0.97 for the ascending and 0.95 for the descending sections), but a lower score for the pulmonary trunk (0.78). Sharkey et al. [25] utilized nnU-Net, tested on internal test set of 100 cases, showing stable results across vessels, with scores of 0.92 for the AAo, 0.91 for the DAo, and 0.93 for the PT. They also evaluated an external test set of 20 cases, where performance remained similar, except for a slightly lower score in the DAo (0.87). Román et al. [26] proposed a new architecture similar to V-Net for PT segmentation, achieving a Dice score of 0.89 on a test set of 91 cases. In summary, U-Net-based deep learning architectures outperform other methods in vascular segmentation but struggle with complex vascular structures like the pulmonary trunk.

Compared to U-Net-based semantic segmentation architectures, AI foundation models like the Segment Anything Model (SAM) have demonstrated improved generalization, efficiency, performance, and accessibility [28]. SAM, developed by Meta AI Research [29], was trained on the largest image segmentation dataset to date, comprising over 1 billion masks across 11 million images. Ma et al. introduced MedSAM, a refined version of SAM specifically designed for medical image segmentation. However, despite these advancements, MedSAM and nnU-Net models achieved similar Dice scores of 0.94 and 0.93 for aorta segmentation [30]. In contrast, our model, tested on a larger dataset of 210 cases, performed consistently well with accuracy rates of 0.95–0.96 across both the aorta and PT, demonstrating competitive results despite utilizing 2D image segmentation and accounting for variable examinations quality. A deterministic algorithm was developed to select the optimal 2D object of the major blood vessels from the candidate pool for measurement. However, this algorithm failed in 3, 8, and 15 cases for the DAo, AAo, and PT, respectively. The primary reason for these failures was inferior or acceptable examination quality of the cases. When our proposed segmentation method was evaluated on a publicly available dataset, it demonstrated comparable performance on both internal and external datasets, achieving a median Dice score of 0.95 (IQR, 0.05) for the AAo on the internal testing dataset and 0.94 (IQR, 0.02) on the external SegThor dataset [21]. Collectively, our method delivers robust and consistent segmentation performance while addressing the limitations of variability in CTPA examination quality and dataset generalization.

Our study provides valuable insights into the measurement of major blood vessels, though it is limited by the constraints of 2D segmentation and measurement. While the majority of the literature focuses on 3D segmentation of the aorta and PT, our 2D approach offers a more efficient and task-specific method for segmenting the DAo, AAo, and PT. This approach provides several practical advantages, including the requirement for only 2D manual annotations for model development, reduced computational power, and lower algorithmic complexity. We observed comparable measurement performance compared to previous studies. On a test dataset of 288 contrast-enhanced chest CT scans, Chettrit et al. reported mean differences between their algorithm and radiologists of −0.94 mm for the AAo diameter and −0.86 mm for the PT diameter [31]. Román et al. [26] measured the mean error of the PT radius by calculating the average Euclidean distance between the segmented surface and the ground truth surface using the Discrete Marching Cubes method, resulting in an error of 1.25 mm. In a prior study, we reported mean differences of −3.20 mm for the PT diameter and 0.51 mm for the AAo diameter between their automated measurements and ground truth [19]. Here, we observed larger discrepancies for the AAo, with a mean difference of 1.78 mm between automated measurements and those of radiologists. However, for the PT diameter, the mean absolute difference between the model’s measurements and those of radiologists was −2.16 mm on our internal dataset, which improved to −1.43 mm on the external FUMPE dataset [22].

There are several limitations to our study. First, the model was initially trained on data collected from a single institution, albeit produced using scanners from a variety of vendors. Second, supplementary structural features, including the right-to-left ventricular diameter ratio [32], contrast reflux into the inferior vena cava [33], and cardiac chamber dimensions [34], can provide valuable prognostic or diagnostic information beyond the diameter of the AAo and PT. Third, due to the unavailability of a publicly accessible dataset, we could not evaluate our proposed model on a large external testing set. Fourth, to validate the generalizability of the proposed model, it should be evaluated on various CT protocols, including coronary CT angiography and venous phase contrast-enhanced CT, in addition to CTPA. Finally, while the proposed method achieved better segmentation results compared to our previous solution, we observed significant variability in IV contrast concentration in the PT and in the diameter of the AAo. This discrepancy is primarily due to the measurement algorithm, which was adapted from our solution but was originally designed for that specific segmentation algorithm. Therefore, to improve accuracy in future studies, a new measurement algorithm needs to be developed specifically for our current segmentation method.

In conclusion, the automated end-to-end deep learning-based segmentation model accurately segmented major blood vessels in the chest cavity, and the adapted vessel measurement algorithm showed strong correlation with radiologists’ measurements. Integrating the highly accurate automated solutions presented in this study into Picture Archiving and Communication Systems (PACS) could be of value for radiologists in routine clinical practice.

## Data Availability

Data generated or analyzed during the study are available from the corresponding author upon reasonable request.

https://competitions.codalab.org/competitions/21145#learn_the_details-dataset

## Abbreviations

AAo: ascending aorta
CADe: computer-aided detection.
CTPA: computed tomography pulmonary angiography
DAo: descending aorta
HU: Hounsfield unit
DL: deep learning
IV: intravenous
nnU-Net: no-new-U-Net
PE: pulmonary embolism
PT: pulmonary trunk

## Acknowledgements

The project was supported by a grant from Analytic Imaging Diagnostics Arena (AIDA) to Tobias Sjöblom. Tomas Fröding and Dimitrios Toumpanakis were financially supported by clinical fellowships from AIDA. Tomas Fröding was financially supported by the Centre for Clinical Research Sörmland, Uppsala University.

---

**Supplementary Table 1.**
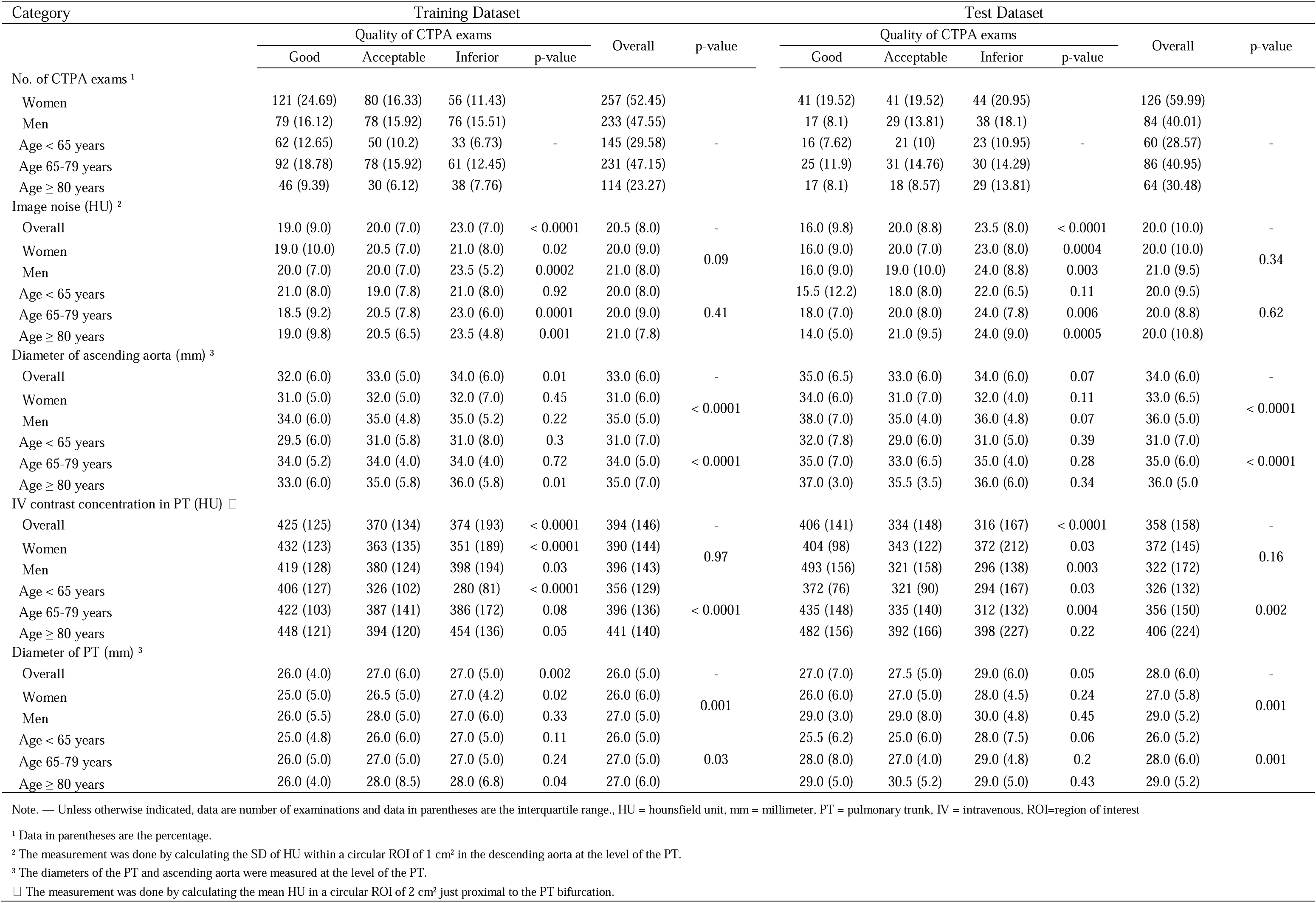
Ground Truth Measurements by Radiologists.

